# Updated population-level estimates of child restraint practices among children aged 0-12 years in Australia, ten years after introduction of age-appropriate restraint use legislation

**DOI:** 10.1101/2022.03.17.22272591

**Authors:** Julie Brown, Bianca Albanese, Catherine Ho, Jane Elkington, Sjaan Koppel, Judith L. Charlton, Jake Olivier, Lisa Keay, Lynne E Bilston

## Abstract

**Objective:** To determine child restraint practices approximately 10 years after introduction of legislation requiring correct use of age-appropriate restraints for all children aged up to 7 years.

**Methods:** A stratified cluster sample was constructed to collect observational data from children aged 0-12 years across the Greater Sydney region of NSW. Methods replicated those used in a similar 2008 study. Population weighted estimates for restraint practices were generated, and logistic regression used to examine associations between restraint type, and child age with correct use accounting for the complex sample.

**Results:** Almost all children were appropriately restrained (99.3%, 95% CI 98.4-100). However, less than half were correctly restrained (No error = 27.3%, 95% CI 10.8-43.8, No serious error = 43.8%, 95% CI 35.0-52.7). For *any error*, the odds of error decreased by 39% per year of age (OR 0.61, 95% 0.46-0.81), and for serious error by 25% per year (OR 0.75 95% CI 0.60-0.93).

**Conclusion:** The findings demonstrate a substantial increase in appropriate child restraint, but no real change in correct use.

**Implications for Public Health:** Given the negative impact incorrect use has on crash protection, continuing high rates of incorrect use may reduce effectiveness of legislative change on injury reduction.

## 1. Introduction

Seat belts and child restraints are effective countermeasures to serious injury and death in car crashes.^1,2^ However, optimal crash protection for child passengers requires the correct use of a restraint that is most appropriate for the child’s size.^1-3^ New legislation was introduced in Australia in 2009/2010 requiring all child passengers aged up to 7 years to correctly use an age-appropriate restraint. Similar child restraint laws exist in many other jurisdictions.^4,5^

In Australia, like many developed countries with long histories of seat belt and child restraint use, the rate of restraint use by children is high. In Australia, restraint use rates have been >90% since the late 1990s,^6^ yet children continue to be seriously and fatally injured in crashes. Population observation studies prior to the introduction of the 2009/2010 legislation indicated around 20% of child passengers were optimally restrained, with approximately one third using restraints inappropriately, another third using their restraint incorrectly, and the remaining one in five incorrectly using an inappropriate type of restraint. ^7^ As the intention of the legislative change was to ensure the proportion of children correctly using age-appropriate restraints increased, we might postulate that child restraint practices in Australia have improved over the last decade.

Pre- and post-observations of children aged 2-5 years in low socioeconomic areas of Sydney immediately before and after introduction of the 2009/2010 age-appropriate restraint use laws found the odds of age-appropriate restraint was almost three times higher, and the odds of correct use was 1.6 times higher post legislation.^8^ However, Koppel et al^9^ found no significant difference in appropriate restraint or correct use in samples of child restraint observations made in the two years before and two years after the introduction of the legislation. While there were substantial differences in the ages of children studied, sampling methods and definitions used for inappropriate and incorrect use between these Australian studies, the contradictions in results reflect similar contradictions in evidence for the impact of child restraint legislation in the literature more broadly.^5^ From a recent systematic review^5^, substantial heterogeneity between different studies makes it difficult to interpret the evidence for the impact of legislation on restraint use, particularly in terms of appropriate and correct restraint use. In addition to heterogeneity issues, there is also a paucity of studies that incorporate gold-standard approaches such as: using observed restraint practices rather than self-reported practices, randomly selected samples rather than self-selected samples, and samples that are population representative rather than convenience samples or focused on one segment of the population. There have been no population-level examinations of observed restraint practices in Australia since our 2008 study or after the introduction of the legislation.

In this study, we aimed to provide an update on child restraint practices in NSW, approximately 10 years after the introduction of age-appropriate legislation. While we do not intend to test the impact of the legislation, we used the same methodological approaches as those in our 2008 study^7^ to allow comparison of the situation before and after the legislation using homogenous and gold-standard observational approaches with stratified random sampling to reduce selection bias. Our original intention was to provide updated restraint practice estimates for the state of NSW; however, our study was impacted by COVID-19 restrictions in 2020 and data collection was limited to the Greater Sydney region. We therefore aim to provide updated estimates for this region only.

## 2. Material and methods

We used a cross-sectional design with a stratified cluster sample constructed to observe restraint practices across a population-representative sample of children aged 0-12 years in the greater Sydney region. Data collection occurred during 2019-2020. Six strata based on Local Government Areas (LGAs), four from inner metropolitan areas and two from outer areas, were randomly selected from all LGAs in the region. The 4:2 ratio was used to select inner and outer areas as corresponding to population distribution across Greater Sydney. Sites where children of different age groups visit regularly were then randomly selected from each LGA: preschool/long daycare centres to capture infants and pre-school aged children, and primary schools to capture school aged children, resulting in 12 data collection clusters.

Participants were children aged 0-12 years and their carer who attended a data collection site during a data collection period. Child-carer dyads were randomly selected as they arrived at the site and invited to participate by on-site researchers before they left their vehicles. Refusals were recorded, and reasons for non-participation noted. If more than one eligible child was in the vehicle, the participant was the one who had last had a birthday. An inability to converse in English, and previous participation in the study were the only exclusion criteria.

The study was approved in August 2018 by the South Eastern Sydney Local Health District Human Research Ethics Committee. Approval to collect data at education sites was also obtained via the NSW School Education Research Applications Process.

### 2.1 Data collection

Data collection methods were the same as that used in the 2008 observational study.^7^ In brief, trained researchers attended the sites over a one-two hour period corresponding with child care/school morning drop-off times. If the driver/carer agreed to participate, the child’s initial observations were recorded as they remained in their restraint within the vehicle. Once the child left the vehicle, a structured interview was conducted with the carer, the height and weight of the child were measured using scales and a portable potentiometer, and a detailed inspection of the restraint installation was conducted. The latter included detail on restraint type, make and model and each possible form of correct/incorrect use of the restraint.

The only difference in data collection method from earlier studies was the addition of a standardised series of photographs used for data quality control.

### 2.2 Variables

Appropriate restraint use was defined using the current legal definitions of appropriate restraint in NSW, Australia (see Table 1). Children were categorised as ‘appropriately’ or ‘inappropriately’ restrained using their age and the restraint type they were using.

**Table 1:** Appropriate restraint definitions based on current restraint use laws in NSW, Australia

| Child age | Restraint type |
| --- | --- |
| 0 – 6 months | Rearward-facing restraint |
| 6 months – <4 years | Rearward- or forward-facing restraint |
| 4 years – <7 years | Forward-facing restraint or booster seat |
| 7+ years | Forward-facing restraint, booster seat or adult seat belt. |

Correct use was defined as use exactly as the manufacturer intended. Each type of error was categorised as an ‘installation error’ or a ‘securing error’, and as ‘minor’ or ‘serious’ depending on the likely degradation in protection or increased risk of injury in a crash introduced by that error.^7^ (see Table 2). This resulted in six correct use outcome variables: (i) any error (whether minor or serious), (ii) any serious error, (iii) any installation error (whether minor or serious) (iv) any serious installation error, (v) any securing error (whether minor or serious), and (vi) any serious securing errors.

**Table 2:**
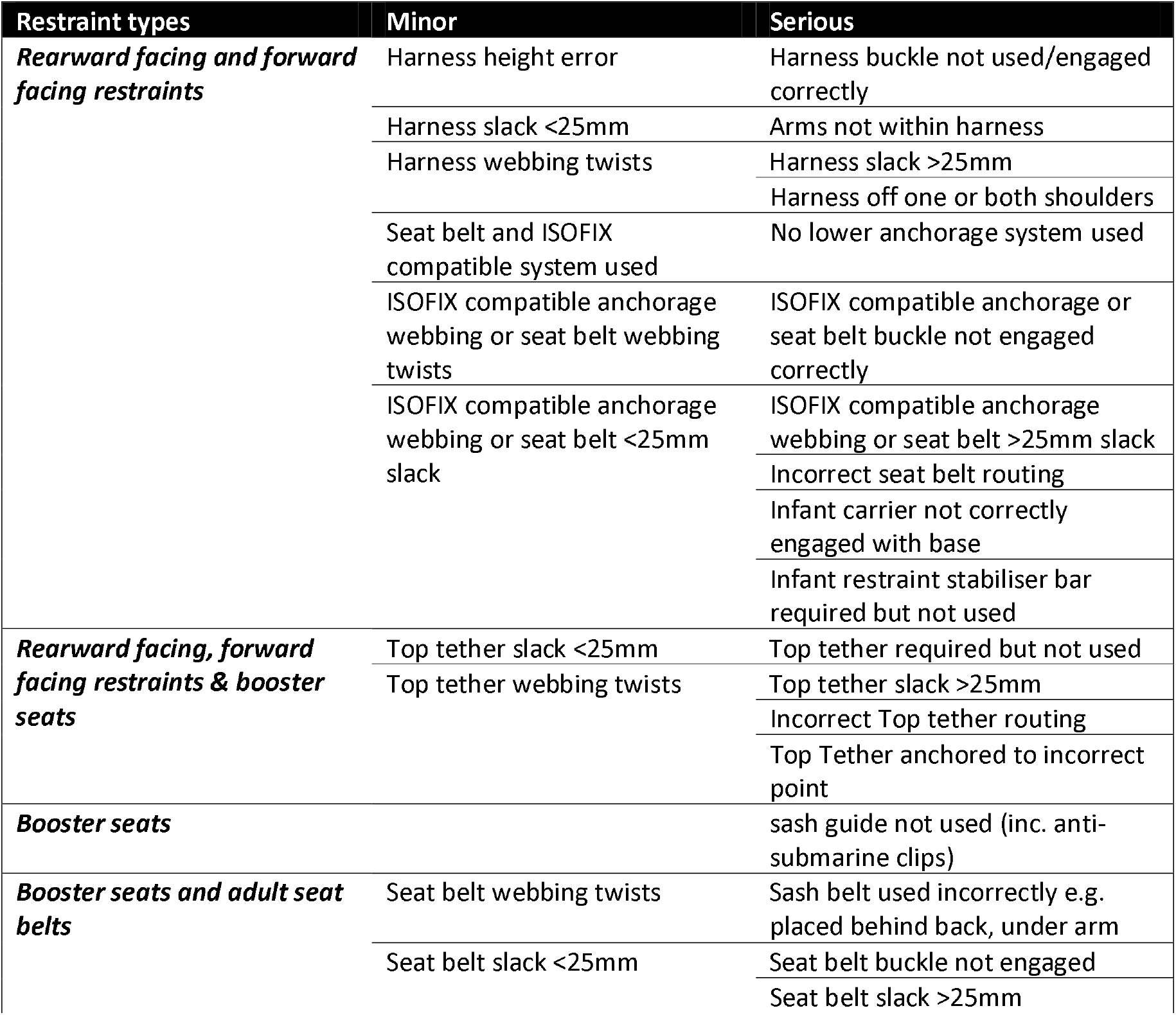
Minor and Serious Errors

Restraints were categorised by type (i.e., rearward-facing, forward-facing, booster seat or adult seat belt) and whether they were convertible or non-convertible restraint types, i.e. designed to be used in more than one mode (e.g., rearward- and forward-facing restraint or forward-facing and a booster seat).

In Australia, rearward and forward facing child restraints can be installed using vehicle seat belt, or ISOFIX compatible connectors. Therefore restraints were also categorised as ‘ISOFIX compatible’ or ‘seatbelt’.

Child age was calculated from date of birth and rounded down to their last birthday.

### 2.3 Data Analysis

All analysis was conducted using SAS Version 9.2 (SAS Institute, 2008). Sample weights were constructed using standard weighting procedures.^10,11^ Post-stratification weighting for age distribution, and under-and over sampling at different sites were used to generate population-level estimates for the Greater Sydney region. The SURVEYFREQ procedure was used to generate population weighted estimates and 95% confidence intervals (CI) for the proportion of children appropriately and correctly restrained, and those using different restraint types. The SURVEYLOGISTIC procedure was used to examine associations between different restraint types, anchorage methods and child age with correct use. The relationship between restraint type and correct use was also examined while controlling for child age, and significant interactions explored through sub-group analysis. Odds ratios (OR) and 95% CI were calculated, and associations assessed as significant when the confidence interval did not pass through 1.

## 3. Results

Data were collected from 213 children aged 0-12 years, sample characteristics are summarised in Table 3. The refusal rate was 40%, with ‘lack of time’ given as the primary reason for refusal. No children were unrestrained, and almost all (99.3%, 95% CI 98.4-100) were using an appropriate restraint as defined by the current legislation. However, fewer children were correctly restrained (i.e., no error = 27.3%, 95% CI 10.8-43.8; no serious error= 43.8%, 95% CI 35.0-52.7). Both installation (any installation error = 53.9%, 95% CI 33.7-74.0, any serious installation error = 36%, 95% CI 24.6-47.4) and securing errors (any securing error = 65%, 95% CI 52.6-77.2, any serious securing error = 40.7%, 95% CI 34.1-47.2) were common.

**Table 3:**
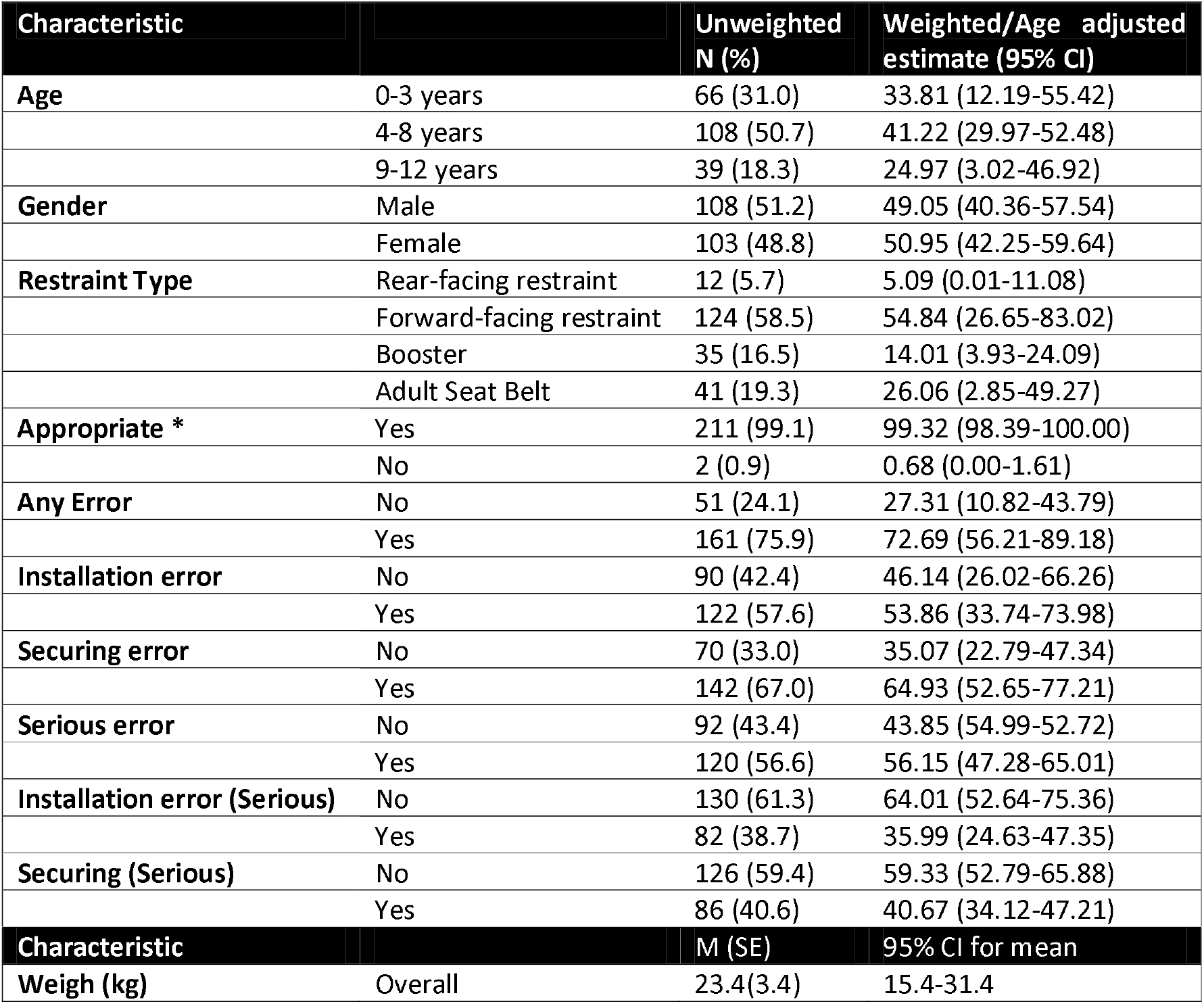
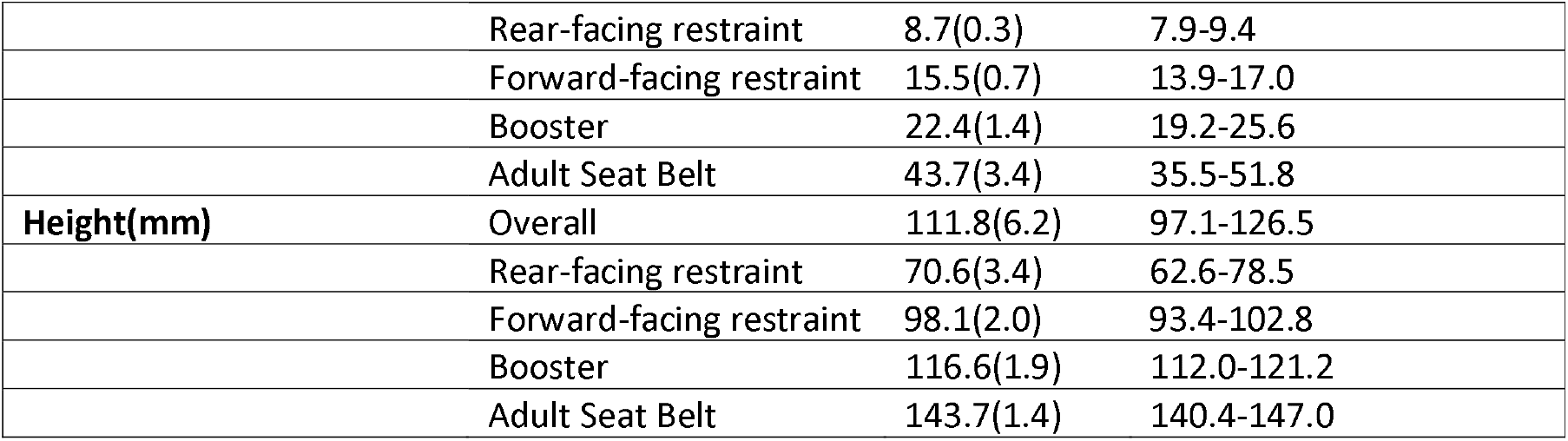
Sample characteristics *NSW legal definition

Across the sample, 5.1% (95% CI 0-11.1) used a rearward-facing restraint, 54.8% (95% CI 26.6-83.0) used a forward-facing restraint, 14.0% (95% CI 26.6-83.0) used a booster seat, and 9.8% (95% CI 2.9-49.3) used an adult seat belt. Most of the rearward- and forward-facing restraints were installed using a seat belt, with 12.5% using an ISOFIX compatible lower anchorage.

Among the children using inappropriate restraint types, all were aged three years and had been prematurely graduated to a booster seat. In addition, all had serious errors in the way the restraint was used with 3/3 failing to use available belt guides resulting in incorrectly positioned belts; one failing to use the top tether, and one failing to engage the seat belt buckle.

Among dedicated restraints (i.e. rear or forward-facing restraints and booster seats), 84% were convertible restraints. There was no association between whether dedicated restraint were single mode or convertible restraints and errors in use (for *any error*, p=0.67; for any *serious error* p=0.84).

Figure 1 illustrates the variation in proportion of children with observed errors by restraint type.

**Figure 1:**
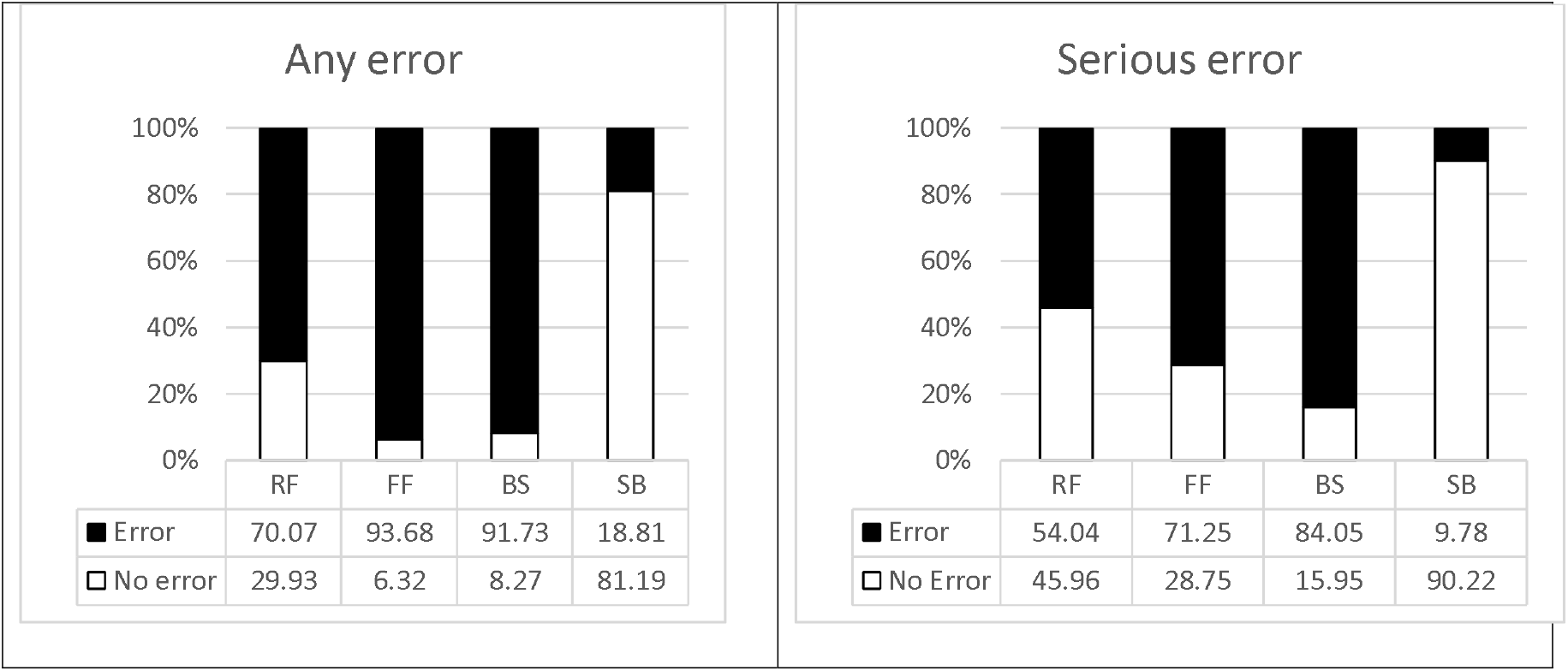
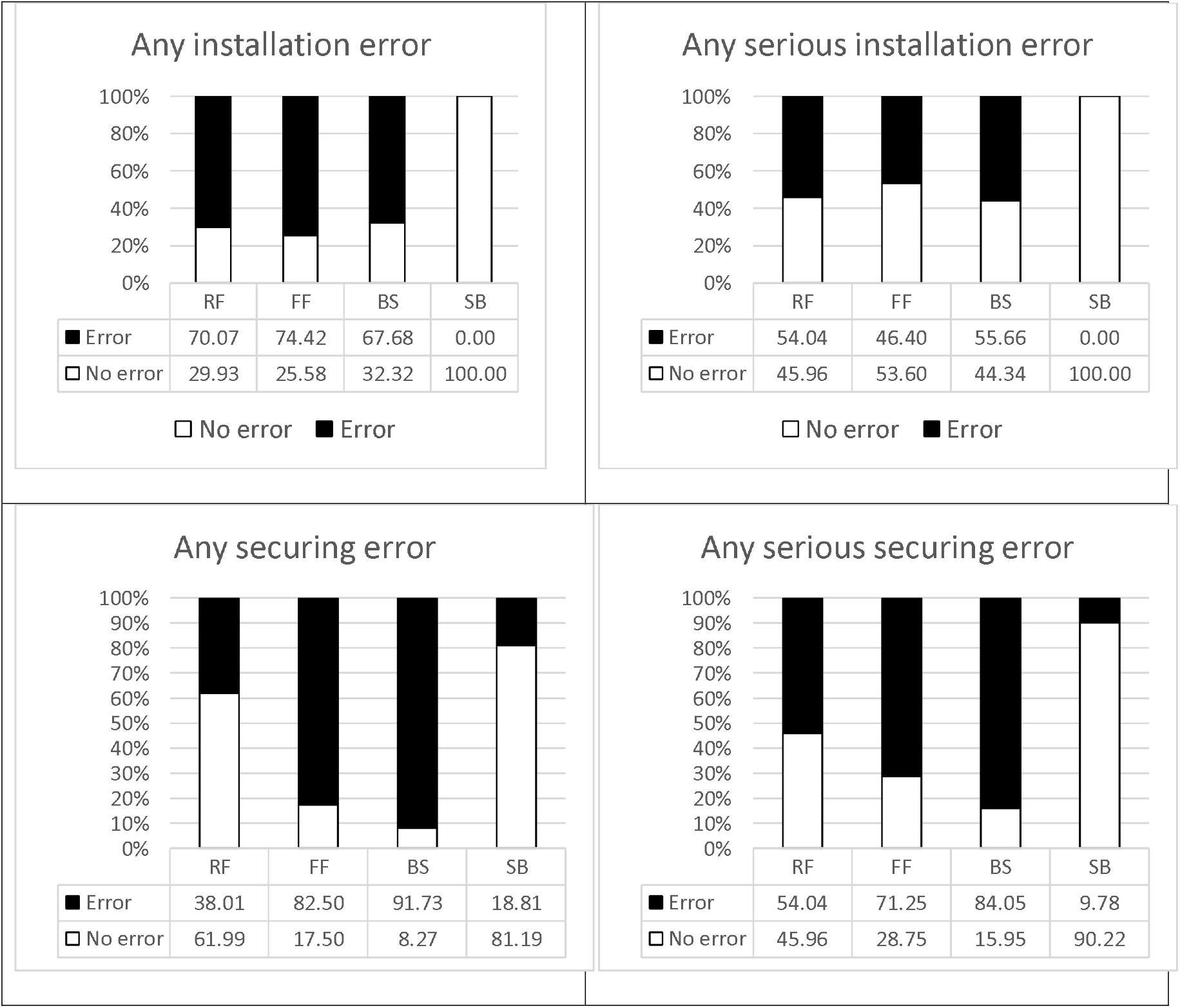
Proportion of errorr, and serious errors by restraint type. RF= Rearward-Facing Restraint, FF=Forward-Facing Restraint, BS=Booster Seat, SB= Seat belt.

Seat belts were used correctly more often than all dedicated child restraint types. Among the dedicated restraints, there were more errors in forward-facing restraints than in rearward-facing restraints (for *any error* in forward-facing restraints compared to rearward facing restraints (OR 6.3, 95% CI 1.18-34.03) but this difference was not significant for serious errors (OR 2.1, 95% CI 0.64-6.88). There were no significant differences in booster seat users compared to rearward-facing restraints (*any error* OR 4.7, 95% CI 0.37-60.7, for *serious errors* OR 4.5, 95% CI 0.84-23.81), or compared to forward-facing restraints (for *any error* OR 0.75, 95% CI 0.17-3.22, for serious errors OR 2.1 95% CI 0.49-9.24).

Examining the types of errors, there were no significant differences in the likelihood of installation errors among the different types of dedicated restraint; however there were significant differences in the odds of securing errors. Forward facing restraint users had over 7 times the odds of securing errors than rearward facing restraints (*any error* OR 7.7, 95% CI 4.9-66.7) but the difference for *serious errors* was not significant (OR 4.8, 95% CI 0.39-58.8). Booster seat users had significantly greater odds for any securing error and serious securing errors than rearward facing restraint users. Booster seat users also had more securing errors than forward facing restraint users but this difference was only significant for serious securing errors (*any error* OR 2.3, 95% CI 0.7-7.8, *serious errors* OR 3.2 95% CI 1.0-10.4).

Figures 2 and 3 illustrate the proportion of children of different ages with and without errors in different types of restraint.

**Figure 2:**
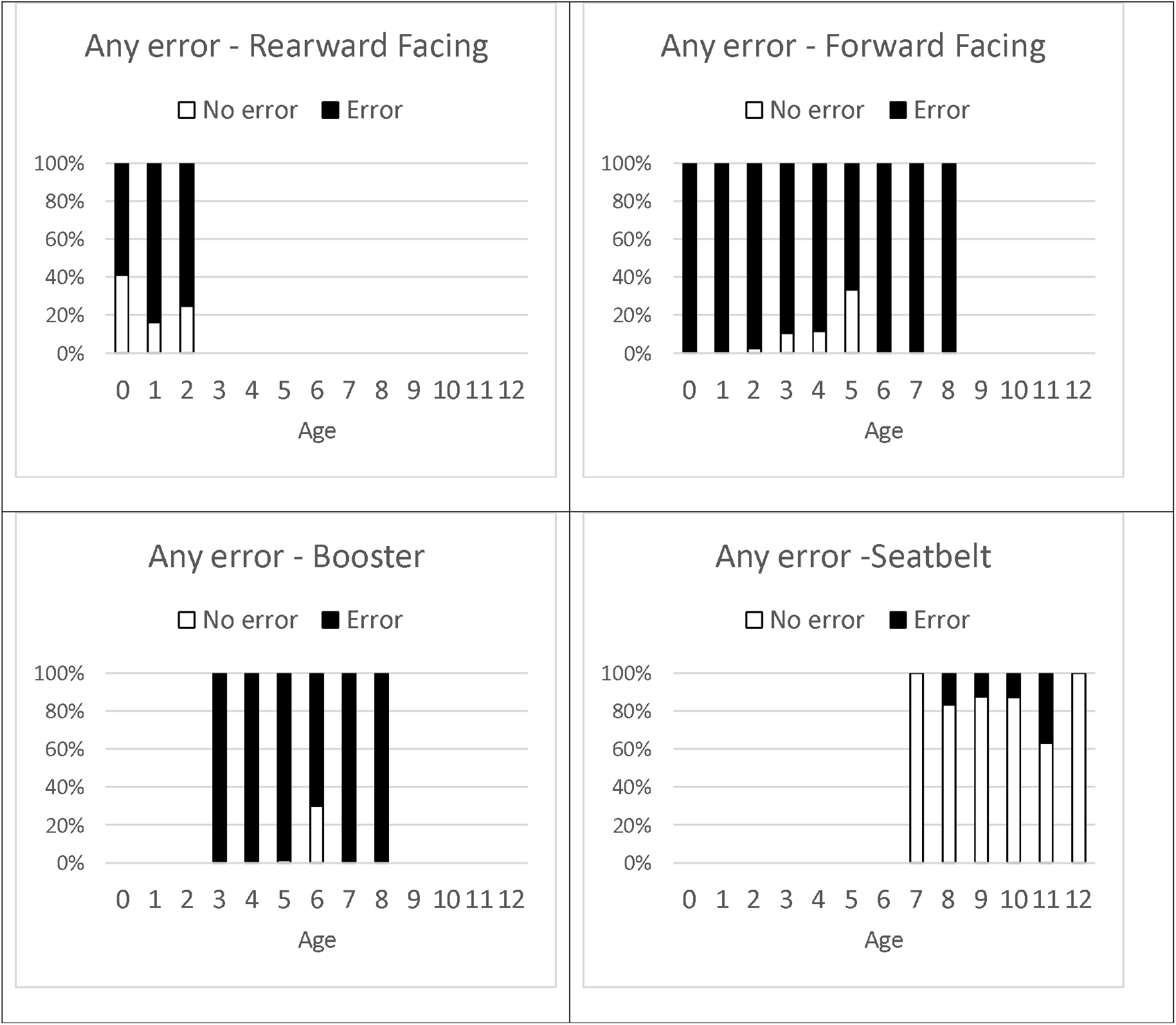
Proportion of children with any error by age for each restraint type.

**Figure 3:**
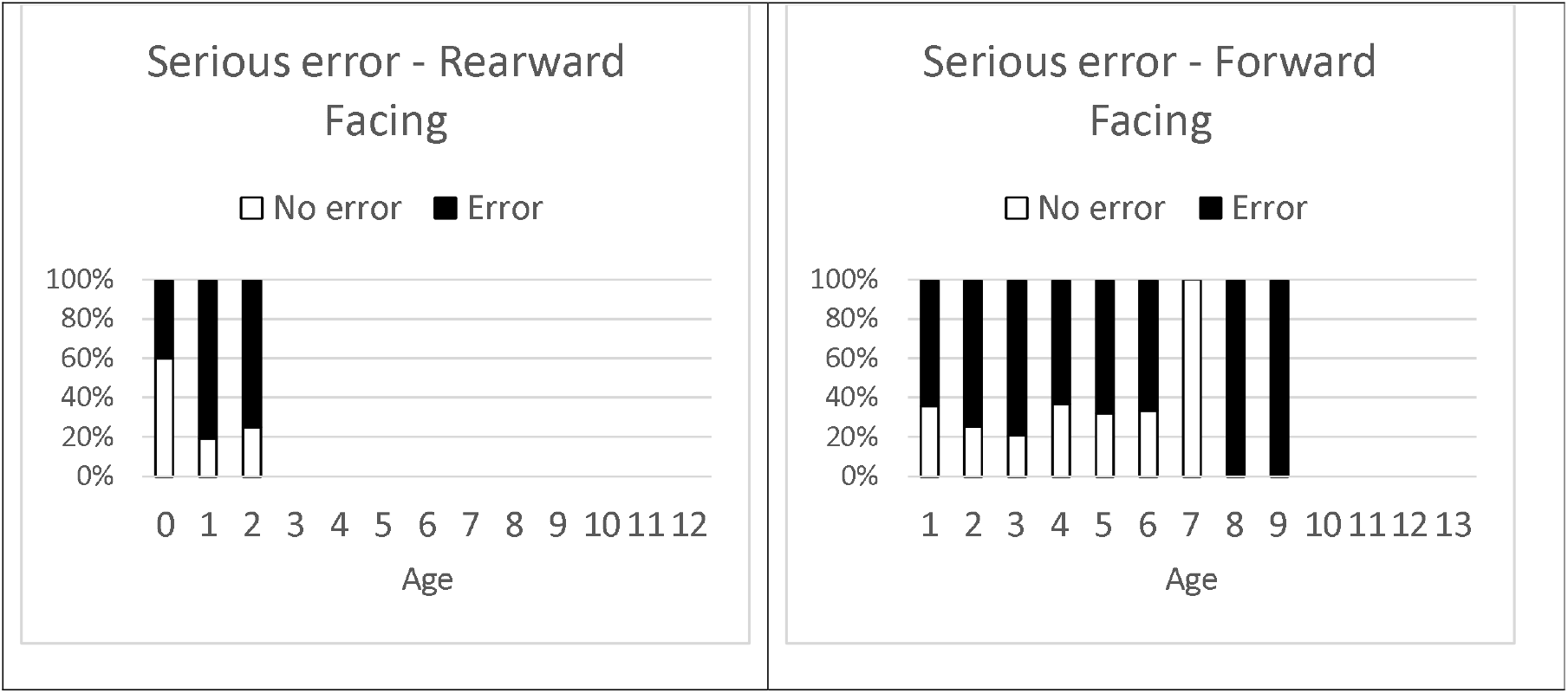
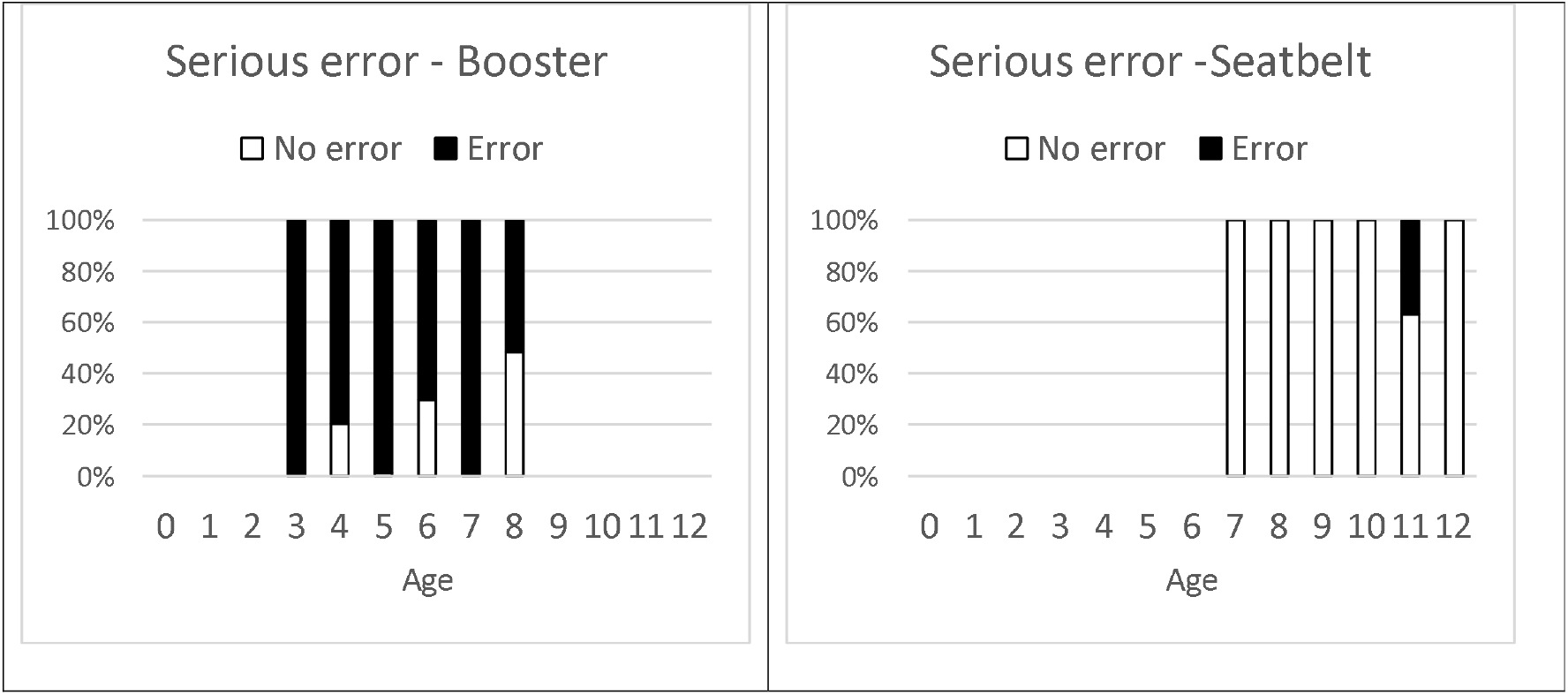
Proportion of children with serious error by age for each restraint type.

The odds of errors decreased with increasing age of the child. For *any error*, the odds decreased by 39% with every year older, and for *serious error* by 25% with every year older (for *any error* OR 0.61, 95% 0.46-0.81; for *any serious error* OR 0.75 95% CI 0.60-0.93). When age was added to the child restraint type models, there was no significant associations between age and errors, or differences between restraint types, for ‘*any error*’ or ‘*serious errors*’. However, there was a significant interaction between age and booster seat use in the ‘any error’ model, and between age and forward-facing restraints in the ‘serious error’ model. This indicates age may have a moderating effect on the likelihood of error by restraint type.

Stratifying by restraint type, age had different relationships with incorrect use. While there was no significant relationship between age and errors in the forward-facing restraints (for *any error*, OR 0.67 95% CI 0.37-1.24/*any serious error* OR 0.91, 95% CI 0.69-1.2), and adult seat belts (for *any error*, OR 1.5 95% CI 0.39-6.09/*any serious error* OR 6.4 95% CI 0.87-46.9), however there was a tendency for the odds of an error to <u>decrease</u> in forward-facing restraints and to <u>increase</u> with increasing age in adult seat belts. For rearward-facing restraints, the odds of errors <u>increased</u> with age *for any serious error* (OR 2.7 95% CI 1.1-6.6), and while there was a similar trend for *any error*, this did not reach significance (OR 2.2, 95% CI 0.86-5.6). For booster seats, the odds of *any error* <u>decreased</u> by 40% with every year increase in age (OR 0.59, 95% CI 0.39-0.89). This relationship was less clear for *any serious error* (OR 0.62, 95% CI 0.17-2.2). Therefore, while overall the odds of errors decreased with increasing age, the relationship between age and errors varied in different restraint types as indicated by the significant interaction in the logistic regression model.

The proportion of errors in rear and forward facing restraints installed using ISOFIX compatible lower anchorage systems were not statistically different (for *any error*, OR 0.29 95% CI 0.05-1.69, for *any serious error* 0.31 95% CI 0.10-1.11, for *any installation error* OR 0.60, 95% CI 0.17-2.05, for *any serious installation error* OR 0.44, 95%CI 0.07-2.62).

## 4. Discussion

This study demonstrates high rates of age-appropriate restraint use among children aged 0-12 years in the greater Sydney region, approximately ten years after introduction of legislation requiring age-appropriate restraint use until age 7. This appears to represent a substantial increase compared to prior to the legislation’s introduction with 99.3% appropriate use in the current study compared to 48.8% in 2008.^7^ However, incorrect restraint use continues to be widespread (51.4% had any error in 2008^7^ and 72.7% in the current study; 38.3% had a serious error in 2008^7^ and 56.2% in this current study).

The results also indicate the uptake of restraints with ISOFIX compatible anchorages is relatively low (∼12% of rearward and forward-facing restraints) compared to North America where the use of these systems is more common than not.^13^ Reasons for this are unknown but might relate to the more recent adoption of ISOFIC compatible restraints and generally older vehicle fleet in Australia. While the lack of significant difference in errors by anchorage method suggests the expected benefit of ISOFIX in reducing installation errors has not been realised, it also possible that user characteristics of those choosing to use or not use this method of anchorage might confound this association. Further exploration of these issues in future work is warranted.

While inappropriate restraint was rare, the characteristics were broadly like those reported in 2008^7^, commonly involving premature graduation to booster seats, and serious errors.^7^ However, there are stark differences between the 2008^7^ and current study in premature graduation to adult seat belts with no observed premature graduation to seat belts in the current study. One important intrinsic difference before and after the introduction of the age-appropriate legislation is how appropriate use is defined. In this study we used the legal age-based definition. Before the legislation was enacted there was no legal definition, and we used restraint type designations set by the product Standard AS/NZS 1754 based on child height and weight to guide appropriate use definitions. To examine the effect of this, the same definitions used in the 2008 study were applied to the current dataset. With those definitions appropriate use in the current sample was slightly lower at 94.2%, (95% CI 87.5-100.0), compared to 99.3%, (95% CI 98.4-100) using the age-based definition, indicating a substantial increase regardless of the definition used. Interestingly, the difference was due to a small number of children aged 7 or 8 years who by the current age-based definitions were appropriately using seat belts but who weighed ≤26kg which was the old upper boundary for appropriate use of booster seats.

It is also important to note that the definitions of appropriate use in this study and in the earlier 2008 study reflect minimum acceptable practices which may be different to best practice. For example, best practice in Australia encourages children be kept in a restraint of a specific type until they reach shoulder height markers fixed to the restraint. With many rearward facing restraints accommodating children up to the age of 2 or 3 years, and some forward-facing restraint accommodating children up to age, best practice would see children remain in these restraint types longer. However, as this depends on the specific make and model which was not always available, it was not possible to compute estimates based on best practice. Similarly, while children aged over 7 years are appropriately restrained by adult seat belts according to Australian law, best practice would be for children to only use an adult seat belt once they can sit with their knees bent over the seat cushion and they achieve a good fit with the adult belt. In a separate analysis using data from the current study^14^ we found that only 40% of children deemed to be appropriately using adult seat belts according to the legal definition actually achieved a good seat belt fit.

Restraint designs change over time, and since the legislation was introduced in Australia there have been two substantial revisions to the Australian Child Restraint Standard (in 2010 and in 2013). It is possible that the increasing complexity of restraint design may be a factor in the higher rates of incorrect use in this current study, as there was an increase in the number of possible compared to the earlier 2008 study.^7^ However, an additional explanation is the shift of children from inappropriate forms of restraint with lower propensity for errors, like seat belts, to more complex dedicated child restraint systems with more opportunity for errors. While the same basic data collection and method of identifying errors was used in this and the earlier 2008 study,^7^ the new quality control measures introduced in this current study (i.e. the secondary check of photographs described in the methods) may also have led to more errors being detected than would have without the secondary review of all cases.

In the current study, we found saw an overall decrease in likelihood of errors with increasing age, consistent with observations made in the earlier study. However, this association was lost when restraint type was controlled for, and we found interactions between age and some restraint types. Our sub-group analysis suggests a more complex relationship between child age, restraint type and incorrect restraint use, but the results of this analysis are limited by a loss of power with smaller sample sizes in the sub-groups. The results from these analyses must therefore be viewed with caution. However, more targeted investigation of this issue in future studies is warranted.

There are several limitations to keep in mind. As data were collected in metropolitan regions, rates of appropriate and correct restraint may be slightly higher than in regional areas, as restraint practices are generally reported to be poorer in rural and regional areas.^5,15^ As this analysis sought to estimate population-level estimates, no attempt was made to estimate differences in restraint use by socioeconomic level and it is possible within the greater metropolitan area, restraint use characteristics might vary from location to location. Estimates from this study may also be biased due to a high refusal rate (∼40%). Ethical conduct of research requires consent for participation, and it is possible that people who are knowingly not using optimal restraint practice may choose not to participate, leading to over-estimations of optimal restraint. Finally, this study did not attempt to examine the impact of vehicle design on correctness of restraint use and the influence of any vehicle design change over time on rates of incorrect restraint use remains unknown.

Strengths of this study include the randomly selected sample and the homogeneity of methods with studies conducted prior to the introduction of the legislation in NSW in 2009/2010. While direct estimates of the effectiveness of the legislation are not possible as other potential influences are not accounted for, use of the same methods allows quantitative comparison of restraint practices before and after the legislative change.

## 5. Conclusion

This study indicates a substantial increase in appropriate use of restraints by child passengers since 2008, likely influenced by the introduction of new legislation requiring age-appropriate restraint use. However, this has not been accompanied by an improvement in correct use of restraints. Given the negative influence of incorrect restraint on the level of protection in a crash, continuing high rates of incorrect use may negatively impact effectiveness of legislation change on reducing injury among child passengers.

## Data Availability

All data produced in the present study are available upon reasonable request to the authors

## 6. Author Contributions

*Julie Brown* Conceptualization, Methodology, Formal Analysis, Writing -Original draft, Supervision, Funding acquisition; *Bianca Albanese Validation*, Investigation, Data curation, Writing – review and editing, Project Administration; *Catherine Ho*, Investigation, Data curation, Writing – review and editing, Project Administration; *Jane Elkington*, Investigation, Data curation, Writing – review and editing, Project Administration; *Sjaan Koppel*, Conceptualisation, Methodology, Writing – review and editing, Funding acquisition; *Judith L. Charlton*, Conceptualisation, Methodology, Writing – review and editing, Funding acquisition: *Jake Olivier*, Methodology, Formal Analysis, Writing – review and editing, Funding acquisition *Lisa Keay*, Conceptualisation, Methodology, Writing – review and editing, Funding acquisition; *Lynne E Bilston*; Conceptualisation, Methodology, Writing – review and editing, Funding acquisition

